# The language network reemerges during recovery from severe traumatic brain injury

**DOI:** 10.1101/2020.01.10.20017004

**Authors:** Brian J. Coffey, Zachary D. Threlkeld, Yelena G. Bodien, Brian L. Edlow

## Abstract

Regaining the ability to express and understand language is a key milestone for patients recovering from severe traumatic brain injury (TBI). However, the neurobiological correlates of language recovery after TBI have not been identified. We explored whether recovery of language in patients with acute severe TBI is associated with functional MRI (fMRI) changes within and outside the canonical language network (i.e. bilateral superior temporal gyri [STG] and inferior frontal gyri [IFG]). We consecutively enrolled 16 adult patients with acute severe TBI and performed fMRI assessment using a spoken language stimulus in the intensive care unit. Eight patients, all of whom recovered language function, returned for follow-up fMRI at median [interquartile range] 220.5 [189-473.5] days post-injury. Sixteen age- and sex-matched healthy subjects also completed the fMRI paradigm. Language function was behaviorally assessed immediately before fMRI using the Coma Recovery Scale-Revised and components of the Confusion Assessment Protocol. At follow-up, patients also completed the California Verbal Learning Test-II. We compared acute and follow-up fMRI responses by calculating mean Z-scores of suprathreshold voxels in bilateral STG and IFG regions-of-interest (ROI). We also performed a whole-brain analysis. Significant longitudinal increases to language stimuli were found in the left STG but not the right STG, left IFG, or right IFG. Whole-brain analysis revealed longitudinal changes in the right supramarginal and middle temporal gyri, regions known to be involved in language processing. Both acute and follow-up fMRI responses in patients were indistinguishable from those of healthy subjects at a stringent statistical threshold of Z ≥ 3.1. At lower statistical thresholds (e.g. Z ≥ 2.1) patients assessed acutely demonstrated decreased fMRI responses in right STG and IFG compared to healthy subjects. Collectively, these results provide initial evidence that responses in bihemispheric language-processing regions of cerebral cortex reemerge with recovery of language function in patients with severe TBI.

## 1. Introduction

Recovery of language is a critical milestone for patients with severe traumatic brain injury (TBI) and an indicator of the transition from unconsciousness to consciousness (Giacino, Kalmar, & Whyte, 2004). Language recovery also predicts better short-term and long-term outcomes for patients with severe TBI (Thibaut, Bodien, Laureys, & Giacino, 2019; Whyte, Cifu, Dikmen, & Temkin, 2001), particularly with respect to social reintegration and return to work (Douglas, Bracy, & Snow, 2016). Despite the key role of language in recovery, the neurobiological mechanisms underlying reemergence of language function in patients with severe TBI are poorly understood. Consequently, there are no early biological markers to identify patients who are likely to recover language function and no interventions to promote this recovery.

It is well established that a left-dominant language network (in right-handed and the majority of left-handed individuals) underlies the three core processes of language function: 1) perception (i.e. identifying sounds as phonetic features, syllables, and words); 2) comprehension (i.e. understanding the meaning of words and sentences); and 3) expression (i.e. using speech, writing, or gesture to express ideas) (Coleman et al., 2007; Friederici, 2002; Price, 2012; Tzourio, Crivello, Mellet, Nkanga-Ngila, & Mazoyer, 1998). This canonical language network is anchored by bilateral nodes in the superior temporal gyrus (STG) and inferior frontal gyrus (IFG) (Demonet et al., 1992; Scott, 2000; Wise et al., 1991). However, the role of the STG, IFG, and other cortical regions in recovery of language function after severe TBI has not been studied longitudinally. Understanding the recovery of language networks after severe TBI may contribute to improved diagnostic and prognostic precision, as well as the development of early interventions targeted at repairing the nodes and connections critical for language perception, comprehension and expression.

We conducted an exploratory, longitudinal study of functional language networks underlying recovery of language in patients admitted to the intensive care unit (ICU) with acute severe TBI. We measured language-evoked fMRI responses in right and left superior temporal gyrus [STG_R_, STG_L_], and right and left inferior frontal gyrus [IFG_R_, IFG_L_]), regions traditionally associated with language function (Mesulam, 1998; Price, 2012). Additionally, we conducted a whole-brain analysis to identify regions outside of this frontotemporal language network related to recovery of language. Finally, to examine whether language recovery is associated with reorganization versus reemergence of language networks, we compared fMRI responses in patients to those in healthy control subjects.

## 2. Materials and Methods

### 2.1 Experimental design

We prospectively and consecutively screened all adult patients with TBI admitted to the ICU at an academic hospital during the 3-year pilot phase of an ongoing trial (ClinicalTrials.gov NCT03504709) approved by the local Institutional Review Board. Inclusion and exclusion criteria have been previously reported (Edlow et al., 2017). Surrogate decision-makers provided written informed consent. Acute fMRI was performed as soon as patients were stable for transport to the MRI scanner, as determined by treating clinicians. Follow-up fMRI and neurobehavioral assessments were performed 6-12 months after injury. Sixteen age- and sex-matched healthy subjects also completed the fMRI paradigm.

### 2.2 Behavioral language assessment

Prior to each fMRI, we assessed patients using the Coma Recovery Scale-Revised (CRS-R) (Giacino et al., 2004), components of the Confusion Assessment Protocol (CAP) (Sherer, Nakase-Thompson, Yablon, & Gontkovsky, 2005b), and the California Verbal Learning Test–II (CVLT-II) (Woods, Delis, Scott, Kramer, & Holdnack, 2006). We defined behavioral recovery of language function as consistent command-following, intelligible expression, functional communication, and the ability to validly complete the CVLT-II.

The Coma Recovery Scale-Revised (CRS-R) is a 23-item, 6-subscale assessment of auditory, visual, motor, verbal, and communication function as well as arousal in patients with disorders of consciousness (DoC) (Giacino et al., 2004). Emergence of volitional cortically-mediated responses (e.g., visual pursuit, object recognition) on CRS-R assessment indicates transition from unconsciousness (i.e, coma or a vegetative state [VS]) to a minimally conscious state (MCS). Return of functional communication or use of common objects is indicative of a post-traumatic confusional state (PTCS). On the CRS-R, grossly intact language function is evidenced by the ability to follow commands, respond to questions, and speak intelligibly (Thibaut et al., 2019). The Confusion Assessment Protocol (CAP) (Sherer, Nakase-Thompson, Yablon, & Gontkovsky, 2005a) is a composite measure of cognition, orientation, and clinical symptoms that measures severity of PTCS. The CAP includes several components that require language comprehension (e.g. a series of four semantically complex questions such as: do you put your shoes on before your socks?) and expression (e.g. verbal responses to prompts). The California Verbal Learning Test-II (CVLT-II) assesses verbal memory and learning. Valid completion of the CVLT-II requires comprehension of the test instructions and intelligible responses to test questions (Woods et al., 2006). Therefore, we used valid completion of the CVLT-II as a proxy for intact basic language comprehension and expression.

### 2.3 fMRI data acquisition

MRI data were acquired with a 32-channel head coil on a 3 Tesla Skyra MRI scanner (Siemens Medical Solutions) located in the Neurosciences ICU. Auditory stimuli were presented to all subjects via MRI-compatible earphones (Newmatic Medical) connected to the scanner’s sound system. The blood oxygen level-dependent (BOLD) functional MRI (fMRI) sequence used the following parameters: echo time=30ms, repetition time=4000ms, in-plane resolution=2.0×2.0mm, slice thickness=2mm, interslice gap=2.5mm, matrix=94×94, field-of-view=192×192mm^2^, 49 slices, 2x GRAPPA acceleration.

Three-dimensional T1-weighted multi-echo magnetization prepared gradient echo (MEMPRAGE) anatomical images were acquired for registration purposes (van der Kouwe, Benner, Salat, & Fischl, 2008): field of view=256×256mm^2^, acquisition matrix=256×256, 176 sagittal slices, 3x GRAPPA acceleration, echo time = 1.69, 3.55, 5.41, and 7.27 ms, repetition time=2530ms, inversion time=1200–1300ms, 1.0 mm^3^ isotropic resolution, flip angle=7°. Patient sedation at the time of acute MRI is reported in Supplemental materials; Table S1. No patients were sedated for follow-up MRI.

The fMRI language paradigm consisted of a block design comprised of two runs, one with a clip from John F. Kennedy’s Inaugural Address played forwards and another with the same clip played backwards. Each run included three 24-s rest blocks and two 24-s stimulation blocks. Prior to the first rest block, 36 s of data were acquired to obtain a stable baseline BOLD signal; these data were excluded from the analysis.

### 2.4 fMRI preprocessing and first-level analysis

Data from the forward language and resting blocks were analyzed. Data from the backward language blocks were not analyzed because our prior study showed that increased STG response to forward versus backward language had a low sensitivity and specificity for identifying patients with behavioral evidence of language function (Edlow et al., 2017).

First-level analysis used the FMRI Expert Analysis Tool (FEAT) (Woolrich, Behrens, Beckmann, Jenkinson, & Smith, 2004; Woolrich, Ripley, Brady, & Smith, 2001) version 6.00 in FSL 5.0.7 (FMRIB Software Library, www.fmrib.ox.ac.uk/fsl) (Smith et al., 2004). Structural and functional volumes were normalized into Montreal Neurological Institute (MNI) space. We applied motion correction using MCFLIRT (Jenkinson, Bannister, Brady, & Smith, 2002), brain extraction using BET (Smith, 2002), and spatial smoothing using a 10mm FWHM Gaussian kernel. To further minimize possible motion-related confounding, we supplemented standard MCFLIRT motion correction with extraction of rotational and translational motion outliers for each dataset using the “fsl_motion_outliers” command. We then included these motion outliers as additional confounder covariates in the general linear model. We contrasted forward language with rest. The resulting Z-statistic images were cluster thresholded (Z ≥ 3.1 and P ≤ 0.05) (Eklund, Nichols, & Knutsson, 2016).

### 2.5 Functional MRI region-of-interest (ROI) analysis

We selected frontotemporal ROIs (STG_L_, STG_R_, IFG_L_, IFG_R_; Supplemental materials; Figure S1) based on prior fMRI studies of language (Adapa, Davis, Stamatakis, Absalom, & Menon, 2014; Davis et al., 2007; Di et al., 2007; Edlow et al., 2017; Liu et al., 2012; Price, 2012) and created ROIs using the Harvard-Oxford Cortical atlas with a probability threshold of 5% (i.e. at least 5% probability that a given voxel is within STG or IFG). We extracted the mean *Z*-score of all suprathreshold voxels (Z <u>≥</u> 3.1) within each ROI using FEATquery in the FMRIB Software Library, (FSL; www.fmrib.ox.ac.uk/fsl). We tested for differences between mean Z-scores in acute and follow-up scans using a Wilcoxon signed-rank test with Bonferroni correction for four comparisons (corrected P significance threshold = 0.0125). We performed statistical analyses with GraphPad Prism 7 (GraphPad; LaJolla, CA).

### 2.6 Functional MRI whole-brain analysis

We conducted whole-brain analyses using the fixed-effects model within FSL (FMRIB’s Local Analysis of Mixed Effects; FLAME) and generated individual and group-level cluster-thresholded Z-statistic maps (Z ≥ 3.1 and P ≤ 0.05). We also compared fMRI responses to language stimuli between patients and healthy subjects at lower statistical thresholds (i.e. Z ≥ 2.3 and Z ≥ 2.1).

### 2.7 Data Sharing

Data processing scripts, stimulus files, and region-of-interest files can be found at: https://github.com/ComaRecoveryLab/LongitudinalLanguagefMRI/. The conditions of our Institutional Review Board ethics approval do not permit public archiving of anonymized study data. Readers seeking access to the data should contact the senior author. No part of the study procedures or analysis was pre-registered prior to the research being conducted.

## 3. Results

### 3.1 Patient demographics and clinical characteristics

We enrolled 16 patients with acute severe TBI and a DoC based on CRS-R and CAP assessment. Eight patients, all of whom recovered language function, returned for follow-up MRI. Injury characteristics and demographics are reported in Table 1.

**Table 1.**
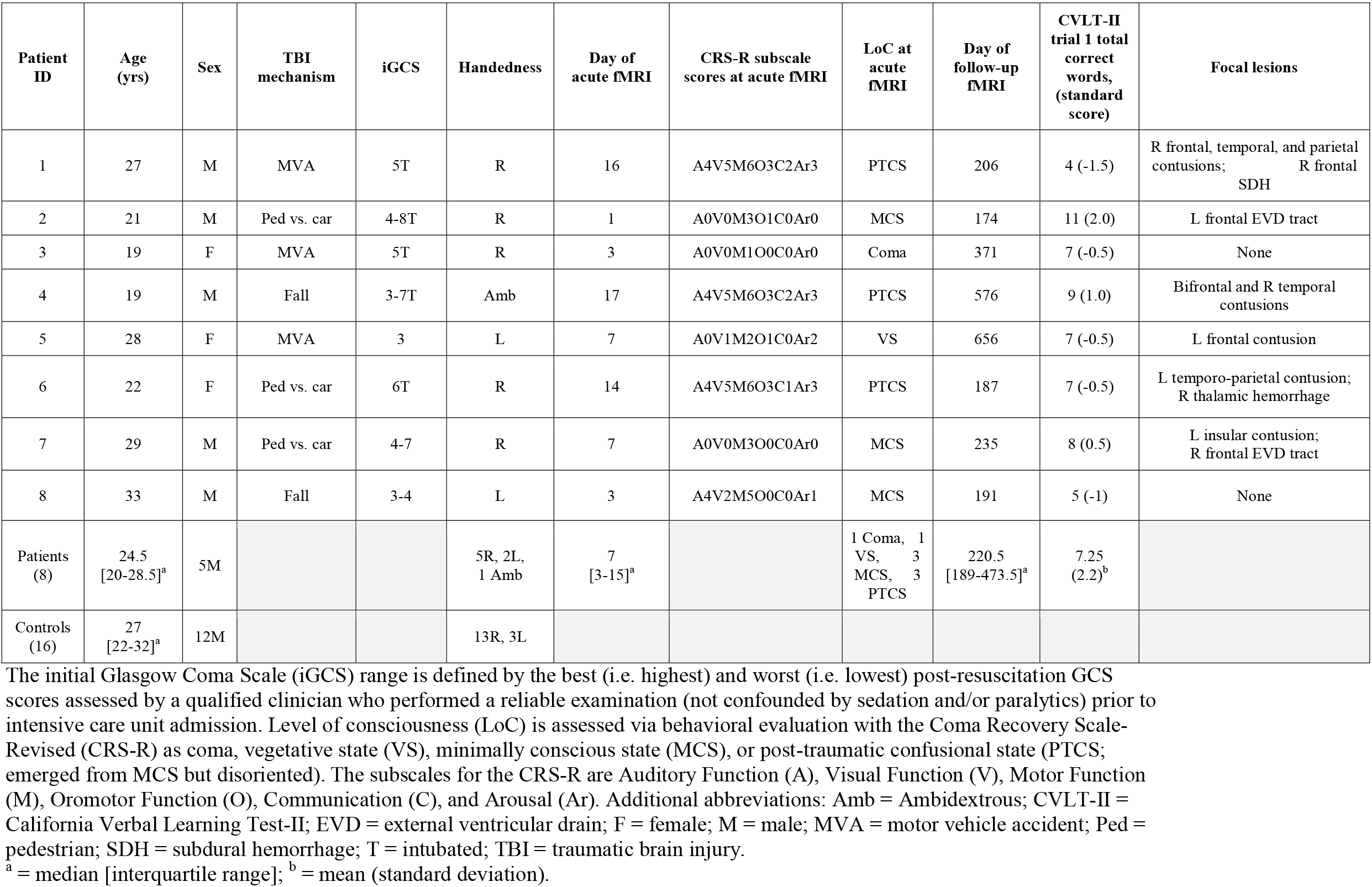
Patient Demographics.

| Patient ID | Age (yrs) | Sex | TBI mechanism | iGCS | Handedness | Day of acute fMRI | CRS-R subscale scores at acute fMRI | LoC at acute fMRI | Day of follow-up fMRI | CVLT-II trial 1 total correct words, (standard score) | Focal lesions |
| --- | --- | --- | --- | --- | --- | --- | --- | --- | --- | --- | --- |
| 1 | 27 | M | MVA | 5T | R | 16 | A4V5M6O3C2Ar3 | PTCS | 206 | 4 (-1.5) | R frontal, temporal, and parietal contusions; R frontal SDH |
| 2 | 21 | M | Ped vs. car | 4-8T | R | 1 | A0V0M3O1C0Ar0 | MCS | 174 | 11 (2.0) | L frontal EVD tract |
| 3 | 19 | F | MVA | 5T | R | 3 | A0V0M1O0C0Ar0 | Coma | 371 | 7 (-0.5) | None |
| 4 | 19 | M | Fall | 3-7T | Amb | 17 | A4V5M6O3C2Ar3 | PTCS | 576 | 9 (1.0) | Bifrontal and R temporal contusions |
| 5 | 28 | F | MVA | 3 | L | 7 | A0V1M2O1C0Ar2 | VS | 656 | 7 (-0.5) | L frontal contusion |
| 6 | 22 | F | Ped vs. car | 6T | R | 14 | A4V5M6O3C1Ar3 | PTCS | 187 | 7 (-0.5) | L temporo-parietal contusion; R thalamic hemorrhage |
| 7 | 29 | M | Ped vs. car | 4-7 | R | 7 | A0V0M3O0C0Ar0 | MCS | 235 | 8 (0.5) | L insular contusion; R frontal EVD tract |
| 8 | 33 | M | Fall | 3-4 | L | 3 | A4V2M5O0C0Ar1 | MCS | 191 | 5 (-1) | None |
| Patients (8) | 24.5 [20-28.5]<sup>a</sup> | 5M |  |  | 5R, 2L, 1 Amb | 7 [3-15]<sup>a</sup> |  | 1 Coma, 1 VS, 3 MCS, 3 PTCS | 220.5 [189-473.5]<sup>a</sup> | 7.25 (2.2)<sup>b</sup> |  |
| Controls (16) | 27 [22-32]<sup>a</sup> | 12M |  |  | 13R, 3L |  |  |  |  |  |  |

The initial Glasgow Coma Scale (iGCS) range is defined by the best (i.e. highest) and worst (i.e. lowest) post-resuscitation GCS scores assessed by a qualified clinician who performed a reliable examination (not confounded by sedation and/or paralytics) prior to intensive care unit admission. Level of consciousness (LoC) is assessed via behavioral evaluation with the Coma Recovery Scale-Revised (CRS-R) as coma, vegetative state (VS), minimally conscious state (MCS), or post-traumatic confusional state (PTCS; emerged from MCS but disoriented). The subscales for the CRS-R are Auditory Function (A), Visual Function (V), Motor Function (M), Oromotor Function (O), Communication (C), and Arousal (Ar). Additional abbreviations: Amb = Ambidextrous; CVLT-II = California Verbal Learning Test-II; EVD = external ventricular drain; F = female; M = male; MVA = motor vehicle accident; Ped = pedestrian; SDH = subdural hemorrhage; T = intubated; TBI = traumatic brain injury.
<sup>a</sup> = median [interquartile range]; <sup>b</sup> = mean (standard deviation).

### 3.2 Standardized behavioral language assessments: Coma Recovery Scale-Revised, Confusion Assessment Protocol, California Verbal Learning Test-II

All patients who returned for follow-up fMRI recovered language function, as demonstrated by the following clinical observations and standardized assessments: 1) intact verbal output, word and sentence repetition, and auditory comprehension on clinical exam, 2) CRS-R total score of 23 (the maximum possible score), 3) correct responses to all semantically complex questions on the CAP and 4) valid completion of the CVLT-II.

On the first trial of the CVLT-II immediate recall test, the mean (SD) total words recalled was 7 (2.2) which is within one SD of the mean, suggesting intact performance. The remaining CVLT-II scores are not reported as they are indicators of verbal memory and learning rather than language function.

### 3.3 ROI-based fMRI responses

Acute fMRI responses observed in the ICU were reported previously (Edlow et al., 2017). Longitudinally, we observed an increase in language-evoked fMRI responses in the STG_L_ (p=0.0117, significant after multiple-comparison correction), increases trending towards significance in the STG_R_ (p=0.0296) and IFG_L_ (p=0.0193), and no change in the IFG_R_ (p=0.4355, Figure 1).

**Fig. 1.**
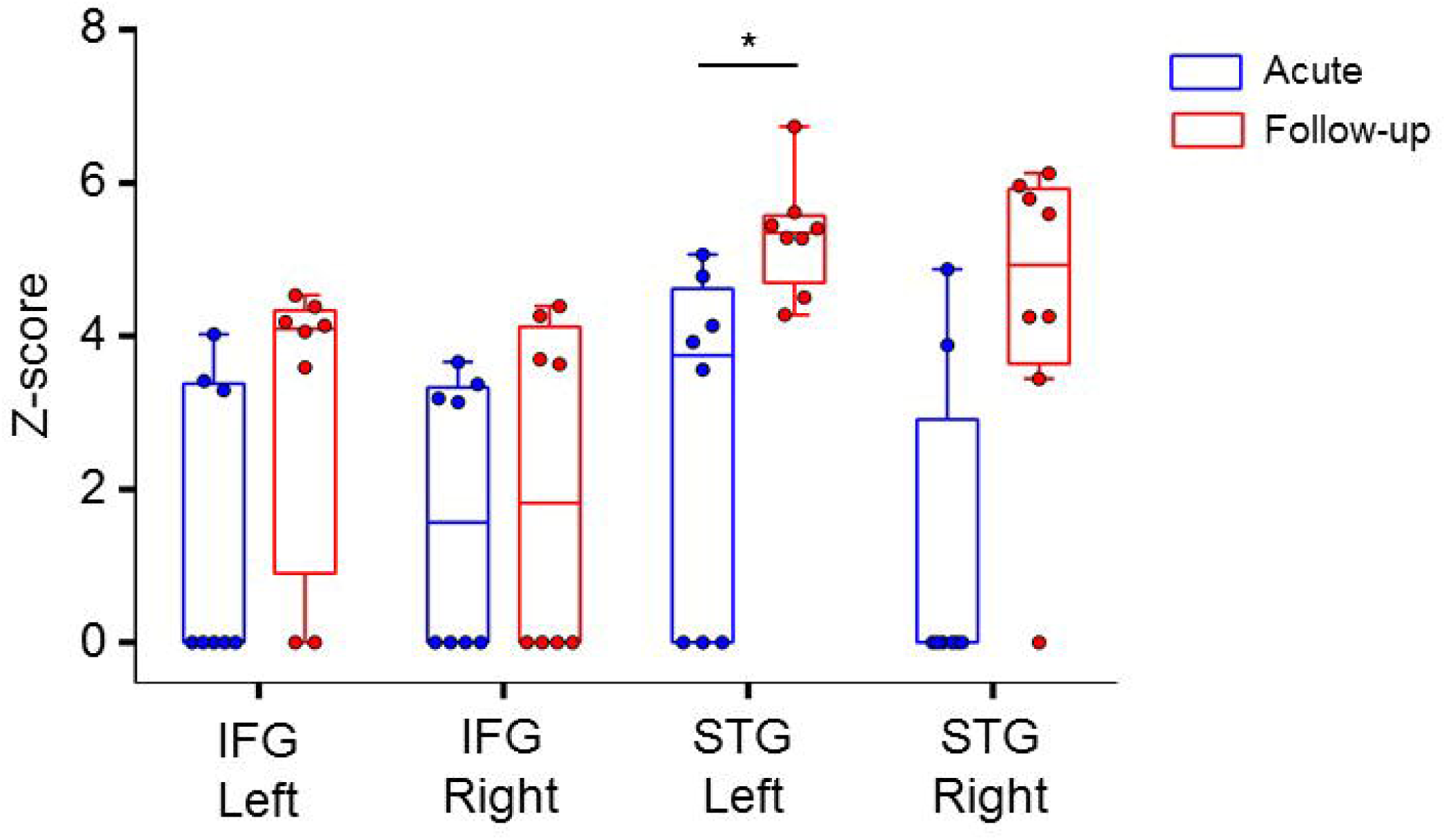
Region-of-interest analysis shows longitudinal increase in left STG responses. The mean *Z*-score of suprathreshold activation in response to the language stimulus is shown stratified by region of interest in acute (blue) and follow-up (red) scans. Box plots illustrate median and interquartile ranges across subjects. There is a significant increase in activation between acute and follow-up scans in the left superior temporal gyrus (STG). We used a Wilcoxon signed-rank test with Bonferroni correction for four comparisons, resulting in an adjusted significance threshold of P<0.0125. *P=0.01; IFG = inferior frontal gyrus.

### 3.4 Whole-brain fMRI responses

In healthy subjects, fMRI responses to spoken language were observed in the bilateral STG and IFG, as well as other cortical and subcortical regions underlying language processing (Supplemental materials; Table S2). Group-level acute patient fMRI responses were limited to language-related regions within the left hemisphere, including the planum temporale in the posterior STG. At follow-up, patients showed responses within the bilateral STG, as well as the right supramarginal, angular and middle temporal gyri. When comparing follow-up to acute patient responses, there were significant longitudinal increases within the right supramarginal and middle temporal gyri (Figure 2), both of which are associated with language processing (Acheson & Hagoort, 2013; Hartwigsen et al., 2010; Visser, Jefferies, Embleton, & Lambon Ralph, 2012; Yue, Zhang, Xu, Shu, & Li, 2013). See Supplemental materials; Table S2 for MNI coordinates of group-level cluster peaks and local maxima.

**Fig. 2.**
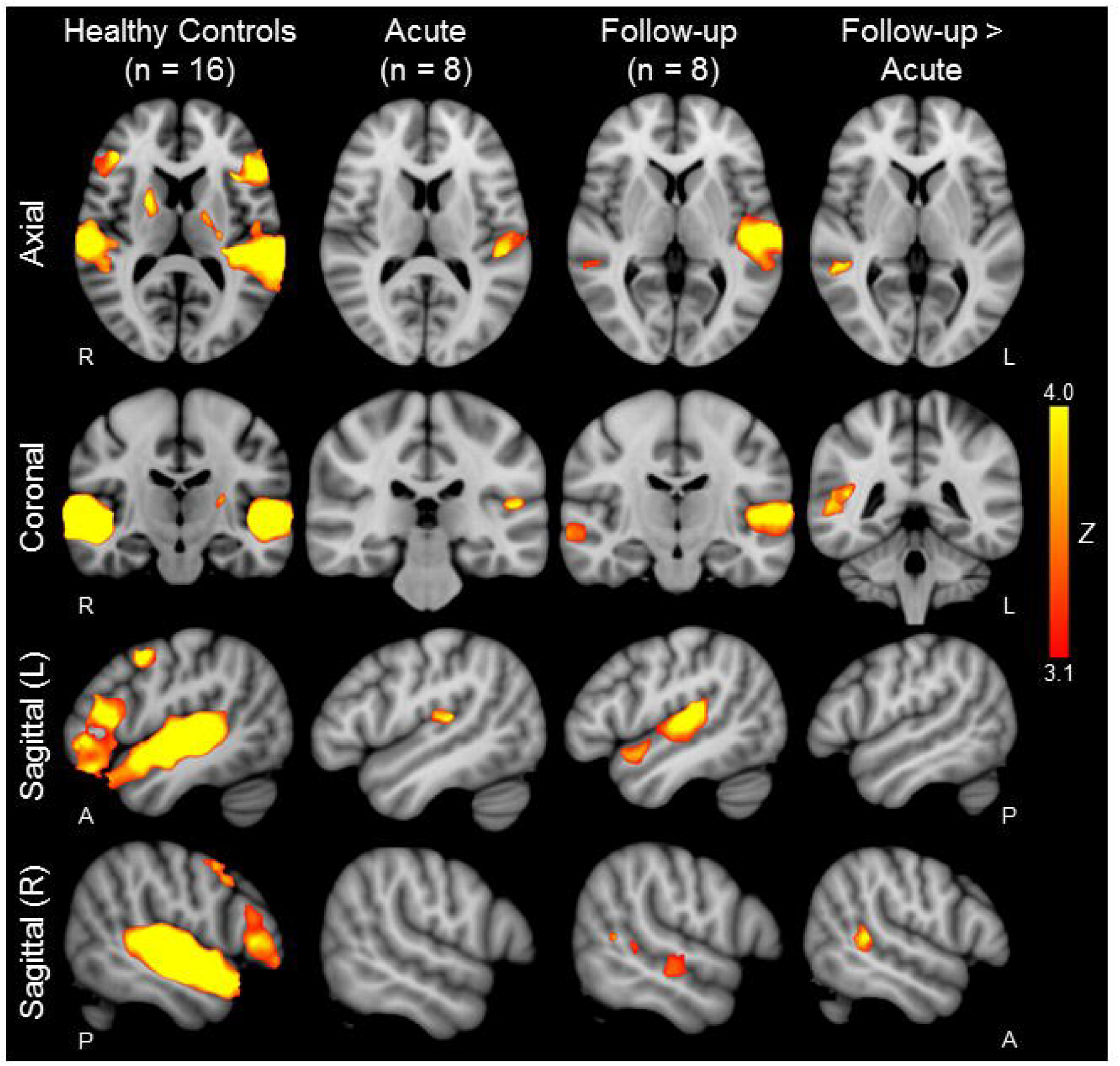
Whole-brain analysis of longitudinal functional MRI responses to speech. Whole-brain group-level functional MRI (fMRI) responses to language stimuli in 16 healthy age and sex-matched controls are shown in the left column. Acute and follow-up whole-brain group-level patient responses to language stimuli are shown in the second and third columns from the left, respectively. Whole-brain group-level longitudinal comparison of follow-up > acute fMRI responses to language stimuli are shown in the right column. *Z*-statistic maps are cluster-thresholded (Z ≥ 3.1 and *P* ≤ 0.05). See Supplementary materials; Table S2 for local maxima coordinates and Harvard-Oxford Cortical Atlas labels. Z = *Z*-score.

### 3.5 Individual Patient Results

Three patients (Patient 3, 4, and 6) did not have STG_L_ activation acutely, but recovered STG_L_ activation by follow-up assessment. Of the six patients (Patients 1, 3, 4, 5, 6, and 8) who lacked STG_R_ activation acutely, five recovered STG_R_ activation by the follow-up MRI. The one patient (Patient 1) who did not show STG_R_ activation at follow-up had significant encephalomalacia in the right temporal lobe due to a massive contusion. IFG_L_ activation was seen in one patient (Patient 1) acutely and 5 patients (Patient 1, 2, 4, 7, and 8) at follow-up. IFG_R_ activation was seen in 0 patients acutely and 3 patients (Patients 2, 4, and 8) at follow-up (Figure 3).

**Fig. 3.**
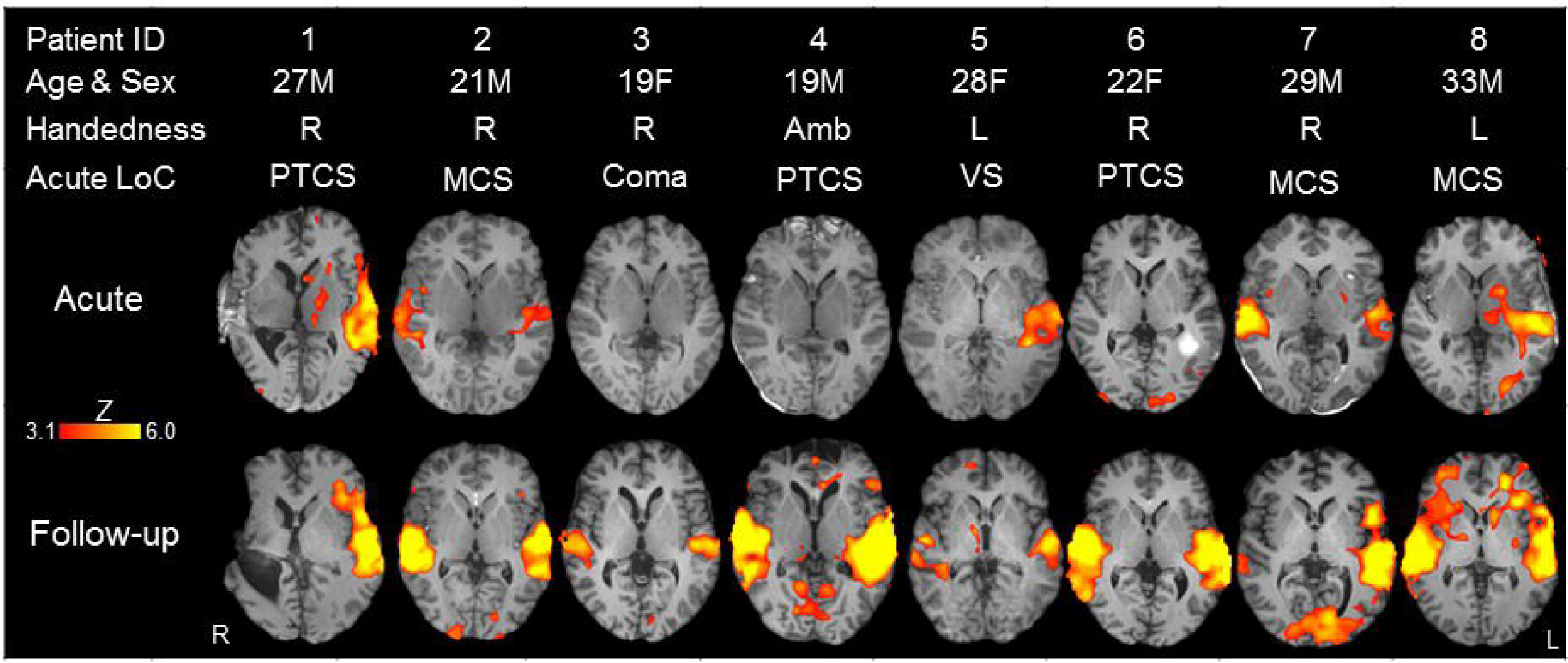
Whole-brain functional MRI responses to language stimuli in each patient at acute (top row) and follow-up (bottom row) time points. *Z*-statistic maps are cluster-thresholded (Z ≥ 3.1 and *P* ≤ 0.05). Abbreviations: Amb = ambidextrous; F = female; L = left; LoC = level of consciousness; M = male; MCS = minimally conscious state; PTCS = post-traumatic confusional state; R = right; VS = vegetative state.

### 3.6 Healthy Control Subjects Compared with Patients

There were no differences in fMRI responses to spoken language between healthy control subjects and patients acutely or at follow-up at the statistical threshold of Z ≥ 3.1. However, when the threshold was decreased to Z ≥ 2.1, patients assessed acutely had reduced activation in right hemispheric regions including the STG_R_ and IFG_R_ (Figure 4 and Supplemental materials; Table S3) while patients at follow-up remained indistinguishable from healthy subjects.

**Fig. 4.**
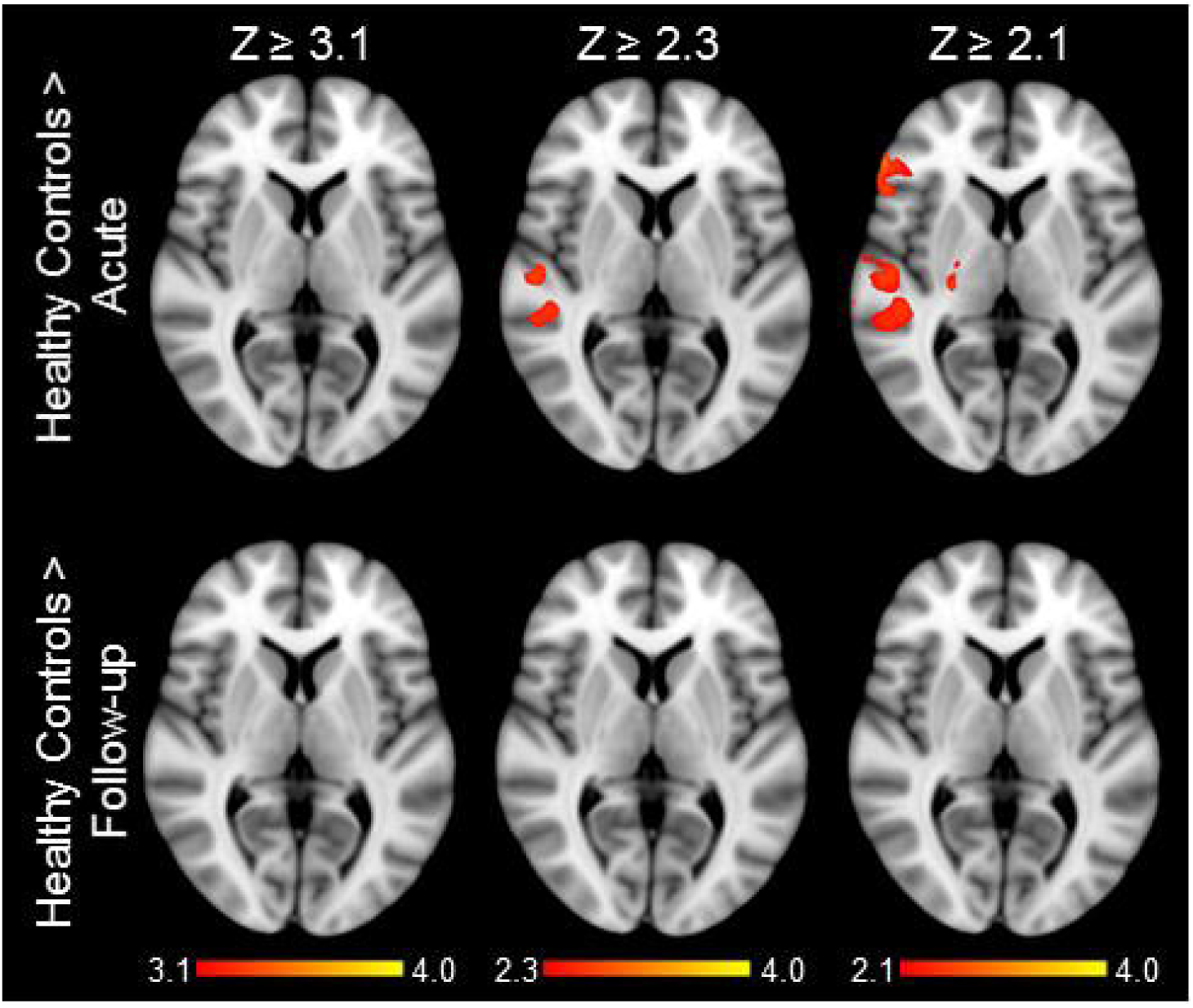
Regions for which fMRI responses to a language stimulus are greater in healthy control subjects than patients acutely (top row) and at follow-up (bottom row) Each column shows *Z*-statistic maps cluster-thresholded at different *Z*-scores (Z ≥ 3.1, Z ≥ 2.3, or Z ≥ 2.1). See Supplementary materials; Table S3 for local maxima coordinates and Harvard-Oxford Cortical Atlas labels. Z = *Z*-score.

### 3.7 Sedation

Six patients received sedative, anxiolytic, or analgesic medications before or during the acute fMRI (Supplemental materials; Table S3). Sedation was not administered for follow-up fMRI.

## 4. Discussion

In patients with severe TBI who recovered language function, we observed longitudinal increases in fMRI responses within the STG_L_, right supramarginal gyrus, and right middle temporal gyrus. Longitudinal changes trended toward increased responses in STG_R_ and IFG_L_ and were absent in IFG_R_, suggesting that not all language cortices are equally involved in recovery of language function after severe TBI. While the mechanistic contributions of the right supramarginal and right middle temporal gyri to language recovery are unknown, the supramarginal gyrus is connected to both the IFG and STG by branches of the arcuate fasciculus (Catani & Mesulam, 2008) and is believed to integrate the meaning of spoken words (Hartwigsen et al., 2010). Similarly, evidence from fMRI and transcranial magnetic stimulation studies suggests that the middle temporal gyrus contributes to comprehension of spoken and written words (Acheson & Hagoort, 2013; Visser et al., 2012; Yue et al., 2013). Collectively, these findings suggest that recovery of language after severe TBI is associated with reemergence of responses within and outside the canonical language network (Jung-Beeman, 2005; Vigneau et al., 2011).

Acutely, we anticipated significantly decreased fMRI responses in patients compared to healthy subjects. Although we did not find this difference at a stringent statistical threshold (Eklund, Nichols, & Knutsson, 2016), patients acutely had decreased right hemispheric cortical responses, including in regions of the language network (STG and IFG). A larger sample size may have revealed this difference at the more stringent threshold. In the absence of pre-injury fMRI data, we cannot differentiate between reorganization (i.e. neuroplasticity) and reemergence of cortical functions underlying language processing. However, fMRI activation maps of patients at follow-up were indistinguishable from healthy control subjects at all statistical thresholds, suggesting that recovery was primarily driven by reemergence, nor reorganization, of language-processing regions.

Our exploratory results, generated from a small but unique longitudinal sample, should be considered in the context of multiple challenges associated with conducting imaging studies in critically ill patients. First, transporting patients from the ICU to the MRI scanner requires a travel ventilator, administration of multiple continuous intravenous infusions, and extraordinary care by nurses, physicians, respiratory therapists and MRI technologists to ensure that lines and tubes do not become dislodged. Second, lying supine in an MRI scanner may exacerbate intracranial hypertension, delaying or even precluding data acquisition during the acute phase of recovery. For patients who survive their ICU hospitalization, returning for follow-up fMRI studies is difficult due to ongoing medical issues, complex transportation needs, and psychosocial factors. Multi-center collaborations will be required to conduct large, rigorous studies that further elucidate the dynamic neurobiological processes underlying recovery of language after severe TBI.

The frequent need for patients to be sedated during the early days of recovery, when the information provided by fMRI may be most useful, is especially problematic for studies of acute severe TBI. The relationship between sedation and fMRI responses is complex and difficult to measure because multiple patient-specific factors, including body mass, tolerance, metabolism, and clearance may alter the effect of a sedative on the BOLD response. However, recent fMRI studies suggest that cortical function is altered by severe brain injury more than by sedation (Bodien, Giacino, et al., 2017; Davis et al., 2007; Edlow et al., 2017; Greicius et al., 2008; Kirsch et al., 2017). Indeed, although six out of eight patients in this study required sedation before and during the acute MRI scan, four out of the six patients requiring sedation demonstrated language-evoked responses in regions underlying language function. Consequently, although the role of sedating medications on cortical responsiveness is not fully understood, the findings in this study are unlikely to be attributable to acute administration of these medications.

Notably, two patients in PTCS, neither of whom could complete CVLT-II testing, did not have language-evoked fMRI responses in the bilateral STG or IFG acutely (Patients 4 and 6; Figure 3). There are three potential explanations for this unexpected finding. First, it is possible that recovery of consciousness and recovery of language function were dissociable in these patients, given that neither patient could complete CVLT-II testing acutely due to inability to comprehend the instructions. Second, the lack of a response in Patient 6 may have been attributable to the effects of lorazepam and haloperidol (though no medications were given to the other patient that did not respond acutely, Patient 4). Finally, a lack of fMRI responses in the bilateral STG or IFG may be due to normal variability in response to the spoken language stimulus. It has been shown that even healthy individuals show variable responses to spoken language stimuli (Otzenberger, Gounot, Marrer, Namer, & Metz-Lutz, 2005).

Because our study did not include patients with persistently altered language function, we are unable to determine whether the fMRI findings are specific to patients who recover language or are generalizable across all patients with acute DoC after severe TBI. Furthermore, although our analyses revealed fMRI changes specific to language cortex, we cannot exclude the possibility that the changes we observed may reflect recovery of consciousness, which co-occurred with recovery of language function. Studies involving serial fMRI and language-specific neurobehavioral assessments are needed to determine the temporal relationship between behavioral language recovery and reemergence of fMRI responses in language cortices.

## Conclusion

In summary, we provide initial evidence that fMRI responses across multiple bihemispheric nodes within the language network reemerge in patients who recover language function after severe TBI. Furthermore, our findings suggest that rather than reorganizing or integrating new nodes, the language network is strengthened and restored following severe TBI. These results provide a foundation for further testing of stimulus-based fMRI as a potential clinical biomarker of language recovery and propose a mechanism of recovery that may be leveraged to develop pharmacologic and rehabilitative interventions.

## Data Availability

https://github.com/ComaRecoveryLab/LongitudinalLanguagefMRI/.

## Acknowledgements

We thank the nursing staff of the Massachusetts General Hospital Neurosciences ICU, Multidisciplinary ICU, and Surgical ICU. We also thank the Massachusetts General Hospital MRI technologists for assistance with data acquisition. We are grateful to the patients and families involved in this study for their participation and support.

